# Face mask use in the Community for Reducing the Spread of COVID-19: a systematic review

**DOI:** 10.1101/2020.08.25.20181651

**Authors:** Daniela Coclite, Antonello Napoletano, Silvia Gianola, Andrea del Monaco, Daniela D’Angelo, Alice Fauci, Laura Iacorossi, Roberto Latina, Giuseppe La Torre, Claudio M. Mastroianni, Cristina Renzi, Greta Castellini, Primiano Iannone

**Author notes:** **Corresponding author**: Silvia Gianola. co-shared role in the authorship, for instance “DC and AN are co-first authors” and “PP and GC are co-last authors”. We must have this symbol and the relative role written on both the authors in order to have the article recognized by the funding source.

## Abstract

**Background:** Evidence is needed on the effectiveness of wearing face masks in the community to prevent SARS-CoV-2 transmission.

**Methods:** Systematic review and meta-analysis to investigate the efficacy and effectiveness of face mask use in a community setting and to predict the effectiveness of wearing a mask. We searched MEDLINE, EMBASE, SCISEARCH, The Cochrane Library and pre-prints from inception to 22 April 2020 without restriction by language. We rated the certainty of evidence according to Cochrane and GRADE approach.

**Findings:** Our search identified 35 studies, including 3 randomised controlled trials (RCTs) (4017 patients), 10 comparative studies (18984 patients), 13 predictive models, 9 laboratory experimental studies. For reducing infection rates, the estimates of cluster-RCTs were in favour of wearing face masks versus no mask, but not at statistically significant levels (adjusted OR 0.90, 95%CI 0.78-1.05). Similar findings were reported in observational studies. Mathematical models indicated an important decrease in mortality when the population mask coverage is near-universal, regardless of mask efficacy. In the best-case scenario, when the mask efficacy is at 95%, the *R*_0_ can fall to 0.99 from an initial value of 16.90. Levels of mask filtration efficiency were heterogeneous, depending on the materials used (surgical mask: 45-97%). One laboratory study suggested a viral load reduction of 0.25 (95%CI 0.09-0.67) in favour of mask versus no mask.

**Interpretation:** The findings of this systematic review and meta-analysis support the use of face masks in a community setting. Robust randomised trials on face mask effectiveness are needed to inform evidence-based policies.

**Funding:** none.

**PROSPERO registration:** CRD42020184963.

**Contribution to the field:** Guidelines by various organizations provide conflicting evidence about the effectiveness of face mask use in the community. We performed a systematic review of the available evidence, including 35 studies, across 41 countries and six continents. Previous systematic reviews on the effectiveness of face mask use mainly focused on healthcare and household setting including only randomized controlled trials and observational studies with most of them of low quality. In our review, we included randomized controlled trials, observational studies, laboratory experimental studies as well as mathematical modelling studies in order to answer different questions and provide quantitative estimates for planning pandemic response efforts.

Our review supports the use of surgical masks in the community for providing protection during the COVID-19 pandemic. However, the infection rate, mortality, spread of transmission (*R*_0_), filtering capacity of masks and viral load reduction are highly dependent on the type of face mask worn and on the adherence of the population wearing masks. Policy makers should promote face mask use in the community.

## INTRODUCTION

Coronavirus disease 2019 (COVID-19) is a new, rapidly emerging infectious disease caused by a novel coronavirus, SARS-CoV-2 (Severe Acute Respiratory Syndrome CoronaVirus-2), which is primarily transmitted via droplets during close unprotected contact with an infector and fomites (1, 2). The virus is genetically similar to the coronaviruses that caused Severe Acute Respiratory Syndrome (SARS) and the Middle East respiratory syndrome (MERS), but SARS-CoV-2 appears to have greater transmissibility and lower pathogenicity than the aforementioned viruses (3). Preliminary estimates of the basic reproduction number (*R*_0_) of SARS-CoV-2, as a metric for transmissibility, range from 2.8 to 5.5, in the absence of intense quarantine and social distancing measures (4). COVID-19 has a higher hospitalization and mortality rate than influenza (5-7) and is spreading in an immune naive population (8). As of 5 July 2020, 11,125,245 people have been infected around the world, deaths are rising steeply up to 528,204 (9). Moreover, there is increasing evidence that people with mild or no symptoms at the pre-symptomatic and early stages of infection can contribute to the spread of COVID-19 (10).

Since there is no effective treatment nor any vaccine for COVID-19, strategies for reducing the burden of the pandemic are focused on non-pharmaceutical interventions for reducing the spread of the infection, such as social-distancing measures, contact-tracing, quarantine, isolation, and the use of face masks in public (11). Public health policies promoting the use face masks in the community, i.e. in public places, can therefore have an important role in controlling the spread of the SARSCoV-2 virus and for COVID-19 lockdown exit strategies (12). The published literature on the efficacy, effectiveness and acceptability of different types of face mask in preventing respiratory infections during epidemics is scarce and conflicting. However, face mask use is increasingly recommended and the potential of this intervention is not well understood (13). National and international health organizations have adopted divergent policies on the subject. Recently, the CDC (Centers for Disease Control and Prevention) and the ECDC (European Center for Disease Prevention and Control) have advocated the use in public places of non-medical face mask (e.g., cloth mask) as a measure for the prevention and/or containment of SARS-CoV-2 infection (10, 14). In areas of significant community-based transmission, where it is difficult to maintain 6-feet social distancing (e.g., grocery stores and pharmacies), CDC recommends wearing cloth face coverings. CDC is additionally advising the use of simple cloth face coverings to slow the spread of the virus and help reduce the transmission of the virus from people who may be infectious without knowing it (14). The World Health Organization (WHO) conditionally recommends face mask use in the community for asymptomatic individuals in severe epidemics or pandemics in order to reduce transmission in the community (15) but it does not recognize its effectiveness in preventing infection (1). Medical and non-medical face masks are used extensively by the general population in Asian countries, such as China, Singapore, South Korea and Japan. Face mask wearing practice has been adopted since the 2003 SARS epidemic in addition to many other response measures and practices, including respiratory etiquette and hand hygiene (10). In Europe, as of 1 April 2020, Lithuania, Austria, Czechia, Slovakia and Bulgaria recommend the use of face masks for persons going out in public (10). Previous systematic reviews on the effectiveness of face mask use mainly focused on healthcare and household setting including only randomized controlled trials (RCTs) with most of them of low quality (16-19). We therefore conducted a systematic review of the existing scientific literature, with randomized trials and observational studies, including modelling and experimental studies, on the effectiveness and efficacy of wearing face masks in the community for reducing the spread of COVID-19 in non-healthcare and non-household setting.

## AIM

The aims of this systematic review (SR) were:

i. to assess the efficacy and effectiveness of using masks in a community setting to reduce the spread of COVID-19 or other similar pandemic; and in particular, to evaluate the effects of using versus not using masks on mortality, infection rate and basic reproduction number (*R*_0_).
ii. to investigate the effect of different filtering capacity of masks used in community settings on the diffusion of the SARS-CoV2.

## METHODS

The systematic review protocol was registered with the International Prospective Register of Systematic Reviews database (PROSPERO identifier: CRD42020184963). The study protocol and preliminary results are publicly available on https://osf.io/uvjgq. We conducted the systematic review following the preferred reporting items for systematic reviews and meta-analyses, the PRISMA statement (20), and the MOOSE guidelines for conducting meta-analysis of observational studies (21).

### Search strategy

We searched for studies on the electronic databases MEDLINE, EMBASE, SCISEARCH and The Cochrane Library from inception to April 22, 2020 using index terms related to face mask use in reducing spread of pandemic infection viruses. Grey literature was interrogated in MedRxiv, Rxiv and bioRxiv databases. We hand searched the reference lists of the included papers. We also incorporated the studies included in any identified relevant systematic reviews. The full search strategy is reported in **Supplementary Appendix 1**.

### Eligibility criteria

According to our PICOS questions (22), the following eligibility criteria without limit of study design were searched:

i. *Population:* general population exposed to SARS-COV-2 infection or other similar virus;
ii. *Intervention and comparators:* any type of mask such as non-medical face mask (i.e., cloth, gauze, tissue), medical face mask (i.e., surgical) and N95 respirators versus no mask;
iii. *Outcomes:* mortality, respiratory infection rate (number of events) and the *R*_0_ of viral respiratory infections; filtering capacity of masks and viral load reduction.
iv. *Setting:* open space community setting for real life situations. Studies assessing the intervention in particular closed cluster setting such as healthcare workers or households were excluded.

### Study selection

Two reviewers independently screened the articles based on the titles, abstracts and full texts. The same two review authors independently retrieved and assessed full reports for potentially relevant studies for inclusion and exclusion according to the above criteria using a predefined electronic spreadsheet. In case of disagreement, consensus was achieved by involving a third independent review author. The reviewers’ decisions and reasons for exclusion were recorded using appropriate reference management software such as EndNote. The study selection process was reported using the flow diagram of the Preferred-Reporting Items for Systematic Reviews and Meta-Analyses (PRISMA) (20).

### Data extraction

Two reviewers independently extracted the study characteristic (e.g., first author, publication year, country, type of virus detected, study design, sample size, settings); for prognostic models, they extracted key characteristics (e.g., factors/predictors, time span, accuracy and performance) and outcomes to be predicted. Disagreements were solved by consensus. A detailed data extraction form was developed prior to the systematic review being performed. In addition, for prediction modelling studies, the Checklist for critical Appraisal and data extraction for systematic Reviews of prediction Modelling Studies (CHARMS) was utilised (23).

### Primary and secondary outcomes

The primary outcomes of this systematic review are the following:

- Mortality rate;
- Respiratory infection rate (measured as event frequency), defined as fever ≥ 37.8 C° with at least 1 respiratory symptom (sore throat, cough, sneezing, runny nose, nasal congestion, headache), with or without laboratory confirmation.
- *R*_0_ of viral respiratory infections; The secondary outcomes were filtering capacity of masks and viral load reduction.

### Data analysis

We examined the efficacy and effectiveness of wearing a mask and the prognostic models available in the literature by study design, setting, and study outcome. The data is summarized in both tabular and narrative formats. As the outcomes were dichotomous, such as respiratory infection, they were analysed as pooled Risk Ratios (RRs), for unadjusted estimates. Adjusted odds ratios from multivariable regression reported in the studies were pooled as adjusted Odds Ratios (aORs). These are summarized using random effects meta-analysis using the DerSimonian and Laird random effects model (24), with heterogeneity calculated from the Mantel-Haenszel model. All summary measures were reported with an accompanying 95% confidence interval. Data analyses were performed using RevMan Software.

### Assessment of study quality

Two independent reviewers appraised the risk of bias. In case of disagreement, a third reviewer was consulted. We used the Cochrane risk of bias tool for randomized controlled trials (25); the Newcastle Ottawa scale for non-randomized studies (26). We planned to use the PROBAST (Prediction model Risk Of Bias Assessment Tool) for Prediction Model Studies (27). However, since we found only quantitative-deterministic models, (statistical) bias was not a suitable measure of model goodness and we analysed the QUAntitative-Deterministic models Risk of Infeasibility Assessment Checklist (QUADRIAC) according to the appropriate guideline (28). We provided more details in **Supplementary Appendix 2**.

### GRADE – quality of the evidence

The Grades of Recommendation, Assessment, Development and Evaluation (GRADE) framework for judging the quality of evidence has been extended to prognosis factor research. Evidence on prognostic models were evaluated by six factors that may decrease quality: (1) phase of investigation; (2) study limitations; (3) inconsistency; (4) indirectness; (5) imprecision; and (6) publication bias; and by two factors that may increase quality: (1) moderate or large effect size; and (2) exposure response gradient (29). Two independent reviewers graded the certainty of the evidence using the GRADE approach. Evidence was presented using GRADE Evidence Profiles developed in the GRADEpro (www.gradepro.org) software.

## RESULTS

### Study selection

A total of 684 records resulted from the searches in the electronic databases (MEDLINE, EMBASE, SCISEARCH) and from pre-prints; eleven additional records were identified through citations. After removing duplicates and excluding irrelevant records according to title, abstract and full text reading, 35 studies met our inclusion criteria for the final inclusion. **Figure 1** shows the flow diagram of the study selection process.

**Figure 1.**
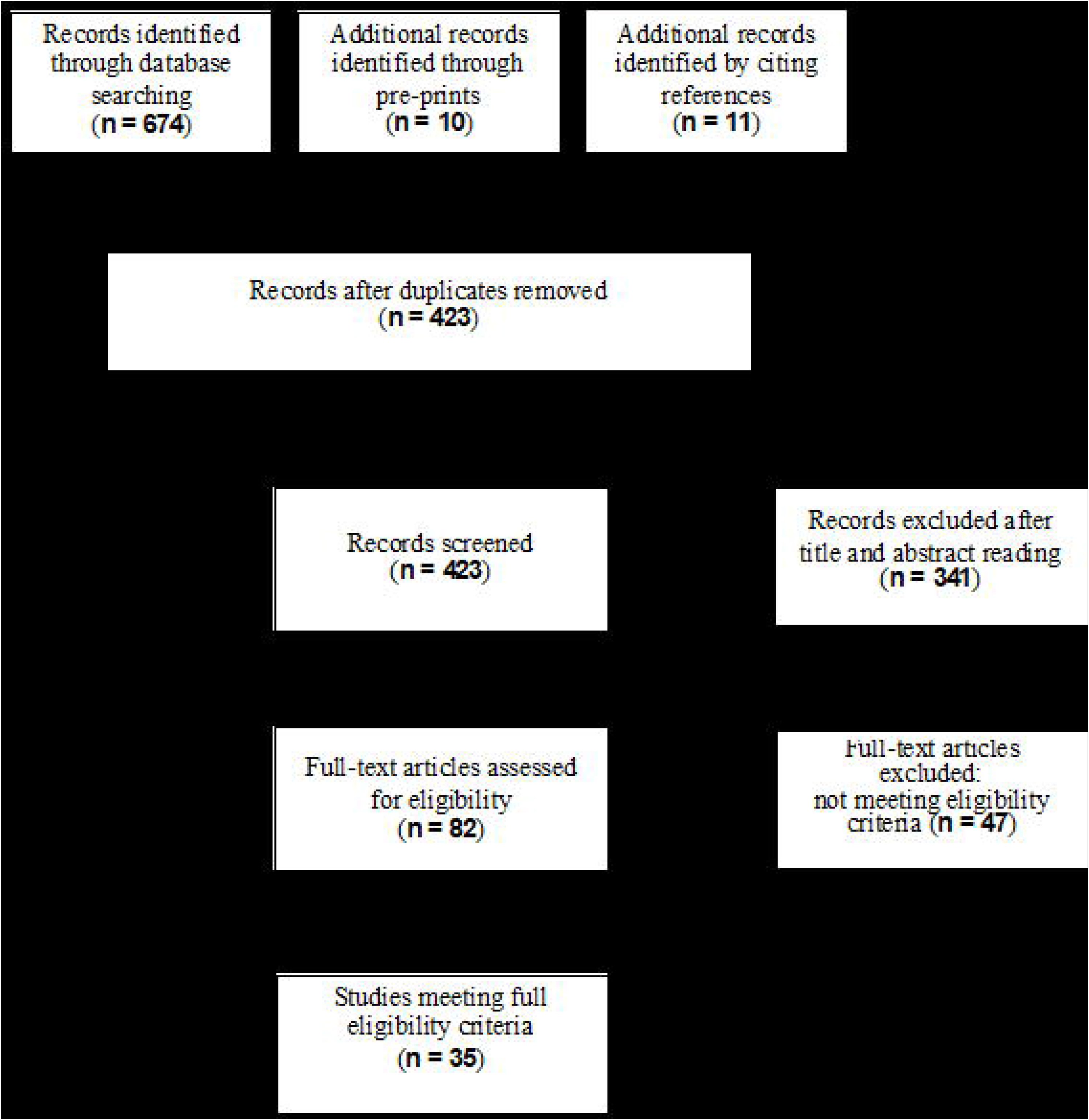
Flow diagram of study selection process

### Description of the included studies

**Table 1** reports characteristics of the included studies. Of the 35 included studies, 3 were cluster-RCTs (30-32), 2 cohort studies (33, 34), 4 were case-control(35-38), 4 cross-sectional (39-42), 13 were quantitative-deterministic predictive models (13, 43-54) and 9 were laboratory experimental studies (55-63).

**Table 1.** - General Characteristics of the included studies

| Study | Study design | Country | Setting | Population size (n) | Disease caused by virus | Adjusted Estimates |
| --- | --- | --- | --- | --- | --- | --- |
| Aiello 2010 | Cluster RCT | USA | University | 930 | H1N1 Influenza | Yes |
| Aiello 2012 | Cluster RCT | USA | University | 816 | H1N1 Influenza | Yes |
| Al-Jasser 2012 | Cohort | Saudi Arabia | Hajj pilgrimage | 1507 | URTI | No |
| Alfelafi 2019 | Cluster RCT | Saudi Arabia | Hajj pilgrimage | 7687 | vRTIs and CRI | Unclear |
| Babak 2020 | SIR-based | Israel | Community | 8 millions | COVID-19 | - |
| Bae 2020 | Controlled Comparison | South Korea | COVID-19 patients | 4 | COVID-19 | - |
| Balaban 2009 | Cohort (pre-post survey) | Saudi Arabia | Hajj pilgrimage | 186 | H1N1 Influenza | No |
| Brienen 2010 | SIR-based | China | Community | not reported | Influenza | - |
| Chen 2008 | SIR-based | Taiwan | Nursery and Primary school | 494 | Influenza | - |
| Choudhry 2006 | Cohort | Saudi Arabia | Hajj pilgrimage | 1066 | ARIs | No |
| Cui 2019 | SIR-based | not reported | Community | 1 million | Influenza H1N1 | - |
| D'Orazio 2020 | Agent-based | Italy | University campus | 5000 | COVID-19 | - |
| Davies 2013 | Controlled Comparison | UK | Healthy volunteers | 21 | Influenza (represented by Bacteriophage MS2 and B atrophaeus) | - |
| De Kai 2020 | SIR-based + Agent-based | 38 selected countries (Asia, Europe and North America) | Community | not reported | COVID-19 | - |
| Deris 2010 | Cross-sectional | Malaysia | Hajj pilgrimage | 387 | ARIs | No |
| Eikenberry 2020 | SIR-based | USA | Community | 1 million | COVID-19 | - |
| Emanian 2013 | Nested case-control | Iran | Hajj pilgrimage | 338 | RTI (all types) | Unclear |
| Guha 2015 | Bench tests | USA | Laboratory | n.a. | Viral infection (Submicron-sized aerosols) | - |
| Kim 2012 | Cross-sectional | South Korea | Primary school | 7448 | H1N1 Influenza | Unclear |
| Lai 2011 | Controlled | China | Laboratory | n.a. | Airborne infections |  |

|  | Comparison (2 manikins) |  |  |  |  |  |
| --- | --- | --- | --- | --- | --- | --- |
| Lau 2004 | Case-control | China | Community | 990 | SARS | Yes |
| Li 2008 | In vivo experiments | Hong Kong | Laboratory | 10 | Viral respiratory infection | - |
| Makison Booth 2013 | Bench test<br>(Head of dummy test) | UK | Laboratory | n.a. | H1N1 and H5N1 influenza<br>(vaccine strain of A-type virus simulation) | - |
| Milton 2013 | In vivo experiment | USA | Laboratory | 37 | Seasonal influenza | - |
| Mniszewski 2015 | Agent-based | USA | Community | 20 millions | Influenza H1N1 | - |
| Ngonghala 2020 | SIR-based | USA | Community | 19.45 millions | COVID-19 | - |
| Rengasamy 2010 | Bench test | USA | Laboratory | n.a. | Influenza | - |
| Tian 2020 | SIR-based | China | Community | 159 | COVID-19 | - |
| Tracht 2010 | SIR-based | USA | Community | 1 million | Influenza H1N1 | - |
| Tracht 2012 | SIR-based | USA | Community | 302 millions | Influenza H1N1 | - |
| Uchida 2017 | Cross-sectional | Japan | Community | 10524 | Influenza | Unclear |
| van der Sande 2008 | Bench test | Netherlands | Laboratory | 61 | Influenza | - |
| Wu 2004 | Case-control | China | Community | 375 | SARS | Unclear |
| Yan 2019 | SIR-based | USA | Community | not reported | Influenza | - |
| Zhang 2013 | Case-control | USA-China | Airplane | 9 | H1N1 Influenza | No |

Of the 13 epidemiological studies (RCTs and observational studies) included in the review, 4 were carried out in a university (30, 31) or school setting (39, 40), 1 on an airplane (37), 6 during mass gatherings (32-34, 38, 41, 42) and 2 in non-specific community settings (35, 36).

As far as the quantitative-deterministic models are concerned, ten studies developed a SIR-based model (11, 13, 43-46, 50-53), two studies developed an agent-based model (47, 48), and one study employed both (54). The whole population was considered in all the studies but one, which was restricted to nursery and primary school children (45). Three out of the thirteen modelling studies (45, 47, 54) considered a closed environment; three models (50, 53, 54) contemplated both inward and outward filtering capacity whereas the others did not make such a distinction; furthermore, only two among the reviewed studies have accounted for a proper usage of facial masks (13, 50). It is worth mentioning that five out of the thirteen studies that have analysed a quantitative-deterministic model have also accounted for the intervention timing (11, 13, 46, 51, 54). Four studies across all the reviewed studies were pre-prints (11, 22, 47, 50).

The laboratory experimental studies were highly heterogeneous in terms of setting/participants: 4 bench test (55, 59, 61, 62), 2 in vivo studies(56, 60) and 3 controlled studies (57, 58, 63).

**Supplementary Appendix 2** lists included and excluded studies.

### Risk of bias of epidemiologic studies and unfeasibility of deterministic models

Focusing on randomized trials, we found high risk of performance and detection bias. However, blinding of participants was not possible due to the nature of the interventions. The included trials were characterized by an overall high quality. Among observational studies the quality ranged from poor to fair for cohort and case-controls studies, whereas it ranged from fair to good for cross-sectional studies. Focusing on mathematical models, we evaluated the unfeasibility of quantitative-deterministic models reporting eight of 13 studies with medium overall risk of infeasibility (2 high and 3 low). **Supplementary Appendix 3** lists the risk of bias of epidemiologic studies and unfeasibility of deterministic models.

### Outcomes

Although no epidemiologic study on wearing face masks in the community for reducing the spread of COVID-19 has been published, a number of studies gave an indirect estimate of the protective efficacy of masks for other viral respiratory infections from agents similar to SARS-CoV2.

#### Mortality rate

##### Deterministic models

Four out of 13 quantitative-deterministic models reported data on mortality (13, 43, 51, 54). Among them, only one study (51) has explicitly provided quantitative data in three scenarios based on different initial values of *R*_0_; however, the time horizon was not specified. Three studies (13, 43, 54) presented graphs depicting the evolution over time of cumulative deaths. Overall the studies point towards a reduction in mortality when the population mask coverage is near-universal, regardless of mask efficacy.

**Table 2** describes the mortality in relation to the initial *R*_0_, type of mask, mask filtration efficacy (%) and adherence of population coverage (%). Summary of findings (SOF) are displayed in **Table 3**.

**Table 2.** Mortality rate in the quantitative-deterministic models.

|  | initial<br>R_0 | type | efficacy % | population coverage % | deaths |
| --- | --- | --- | --- | --- | --- |
| Tracht 2012 | 1.25 | N95 | 20 | 0 | 286,236 |
|  |  |  |  | 10 | 26.445 |
|  |  |  |  | 25 | 2.468 |
|  |  |  |  | 50 | 1.730 |
|  | 1.30 | N95 | 20 | 0 | 327.270 |
|  |  |  |  | 10 | 56.280 |
|  |  |  |  | 25 | 8.301 |
|  |  |  |  | 50 | 2.027 |
|  | 1.35 | N95 | 20 | 0 | 356.462 |
|  |  |  |  | 10 | 84.131 |
|  |  |  |  | 25 | 8.301 |
|  |  |  |  | 50 | 2.573 |
| Eikenberry 2020 | extraction not possible - only graphs |  |  |  |  |
| De Kai 2020 | extraction not possible |  |  | 0<br>50<br>80 | not reported<br>240.000<br>60.000 |
| Babak 2020 | 2.2 | not reported | 8<br>16 | near-universal<br>(% not reported) | extraction not possible |
|  | 1.3 | not reported | 8<br>16 | near-universal<br>(% not reported) | extraction not possible |

**Table 3.**
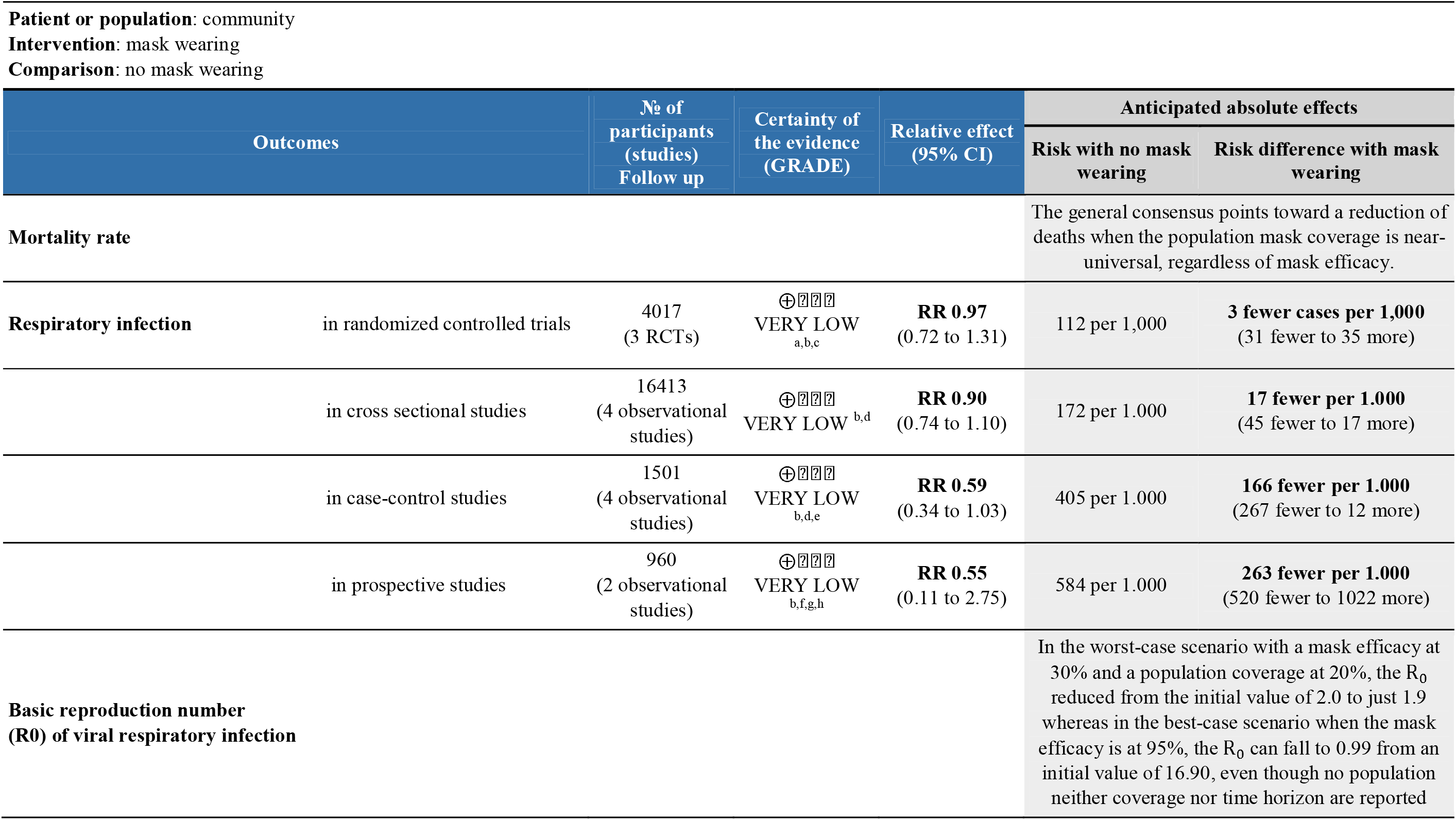
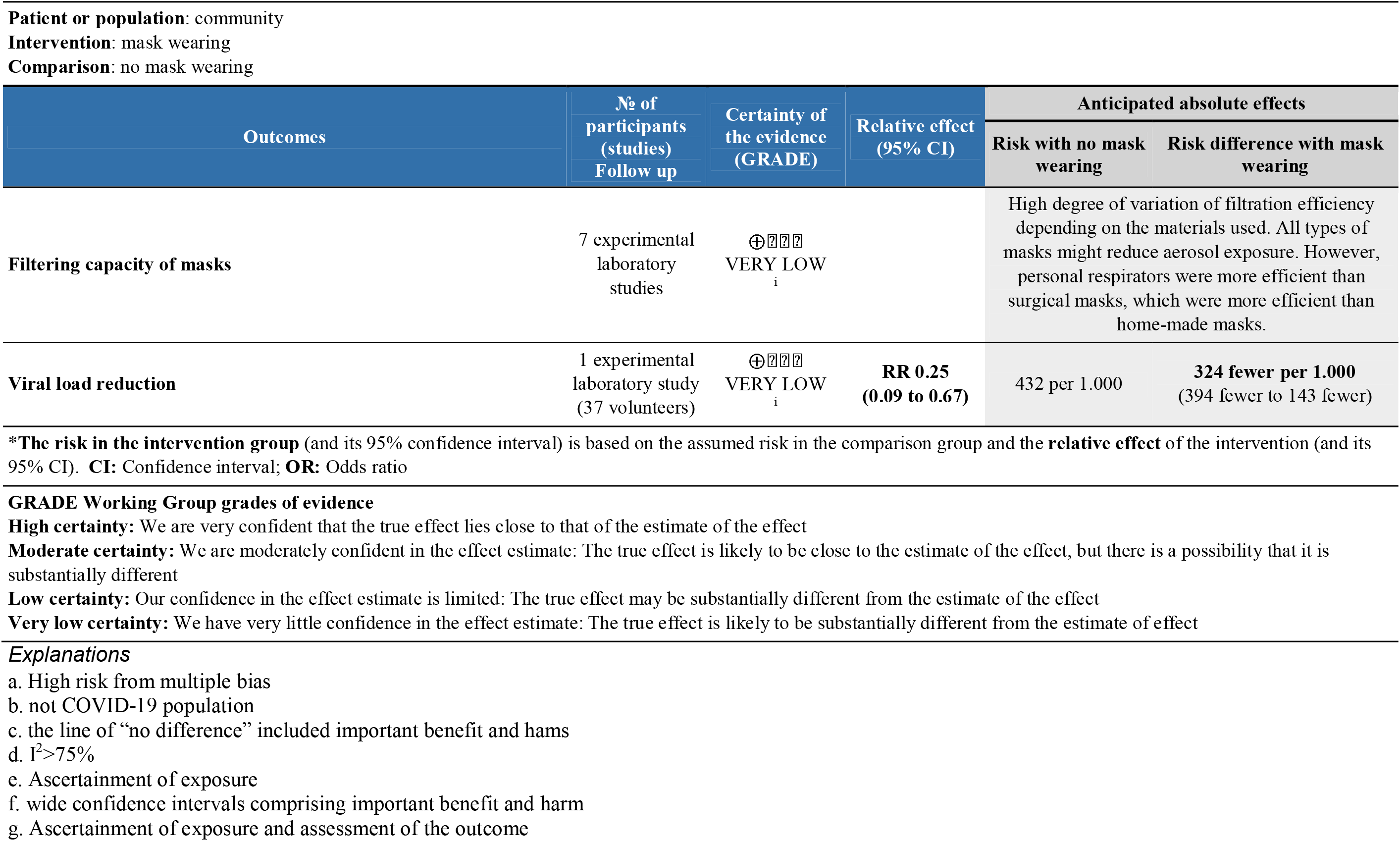
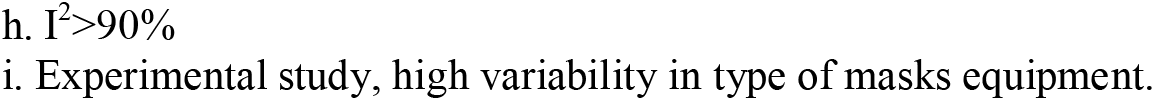
Summary of findings. Wearing a mask compared to no mask in a community setting.

#### Respiratory infection rate

##### RCT

The overall findings were similar between adjusted and unadjusted estimates. With very low quality of the evidence (**Table 3**), in the unadjusted data, three cluster-RCTs (30-32) have reported a small non-significant reduction in the risk of respiratory infections (Figure 2a, RR 0.97, 95%CI 0.72-1.31, I^2^= 62%). The adjusted estimates of two, out of three, cluster-RCTs (30, 31) confirmed the reduction with high consistency, even if not at statistically significant levels (Figure 2b, aOR 0.90, 95%CI 0.78-1.05, I^2^= 0%).

**Figure 2a.**
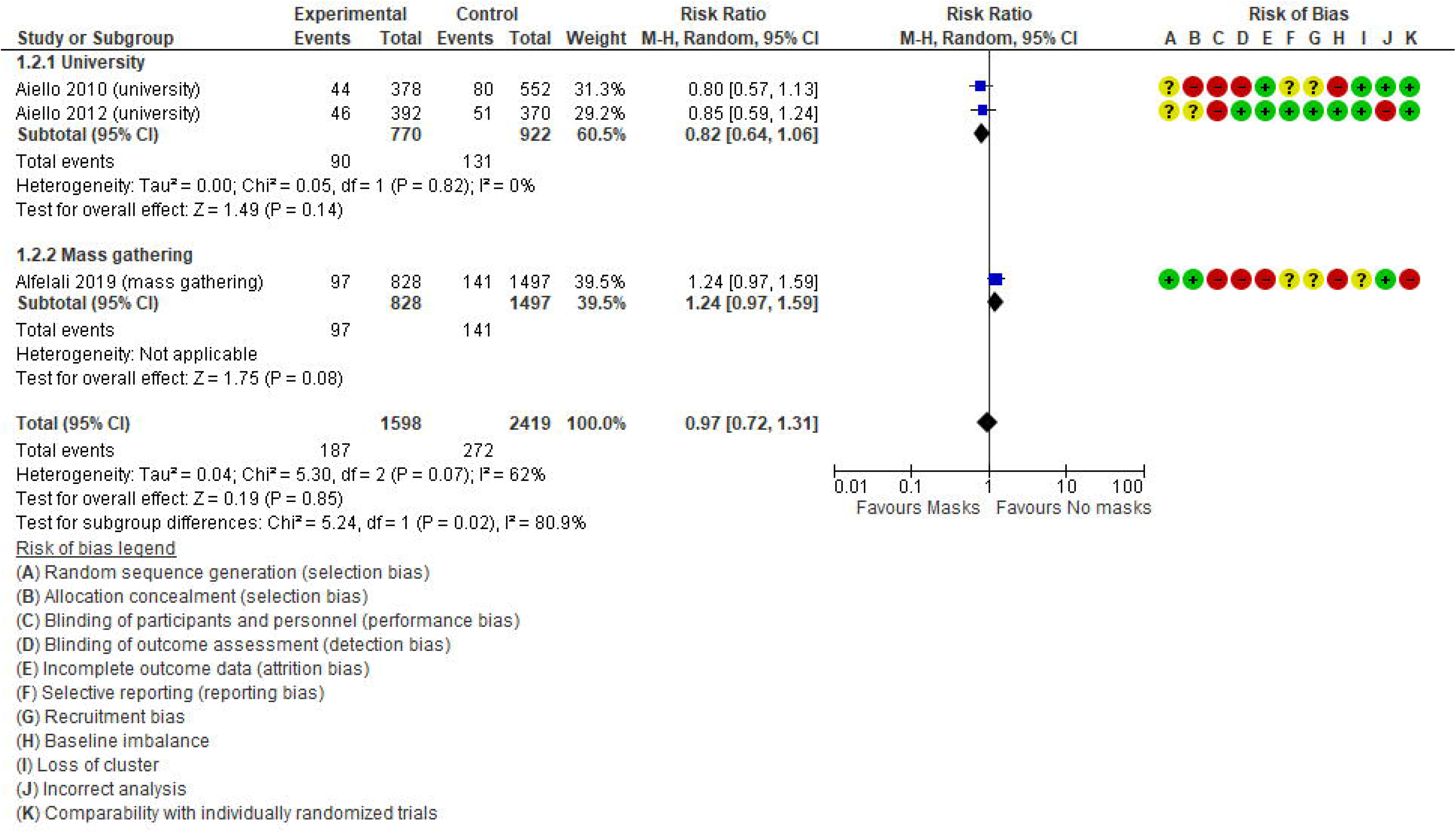
Unadjusted forest plot of respiratory infection rate and risk ratios in RCTs

**Figure 2b.**
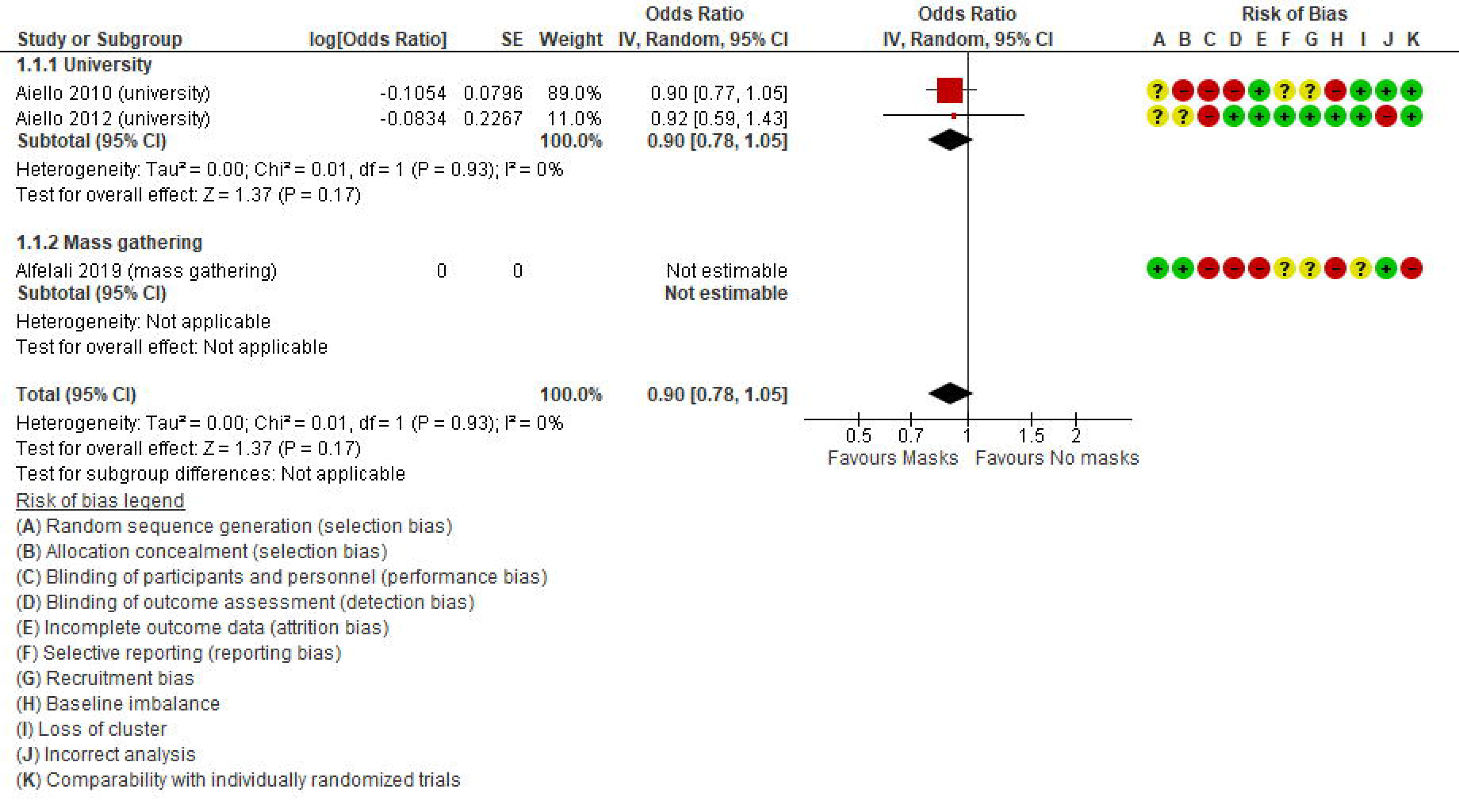
aORs forest plot of respiratory infection rate in RCTs

##### Observational studies

In total, ten observational studies were identified (33-42), among which one study reported adjusted data in relation to the outcome of interest (36). Thus, the meta-analysis was reported only for unadjusted estimates. The level of certainty of the evidence in all observational studies was very low (**Table 3**), with no statistically significant effect; the overall effect was very imprecise across all cross sectional studies (4 studies, OR 0.90, 95%CI 0.74-1.10, I^2^= 74%) (**Figure 3**, Comparison 1.3.1) (39-42), case-control (4 studies, OR 0.59, 95%CI 0.34-1.03, I^2^= 78%) (**Figure 3**, Comparison 1.3.2)(35-38) and prospective cohort studies (2 studies, OR 0.55, 95%CI 0.11-2.75, I^2^= 97%) (**Figure 3**, Comparison 1.3.3)(33, 34).

**Figure 3.**
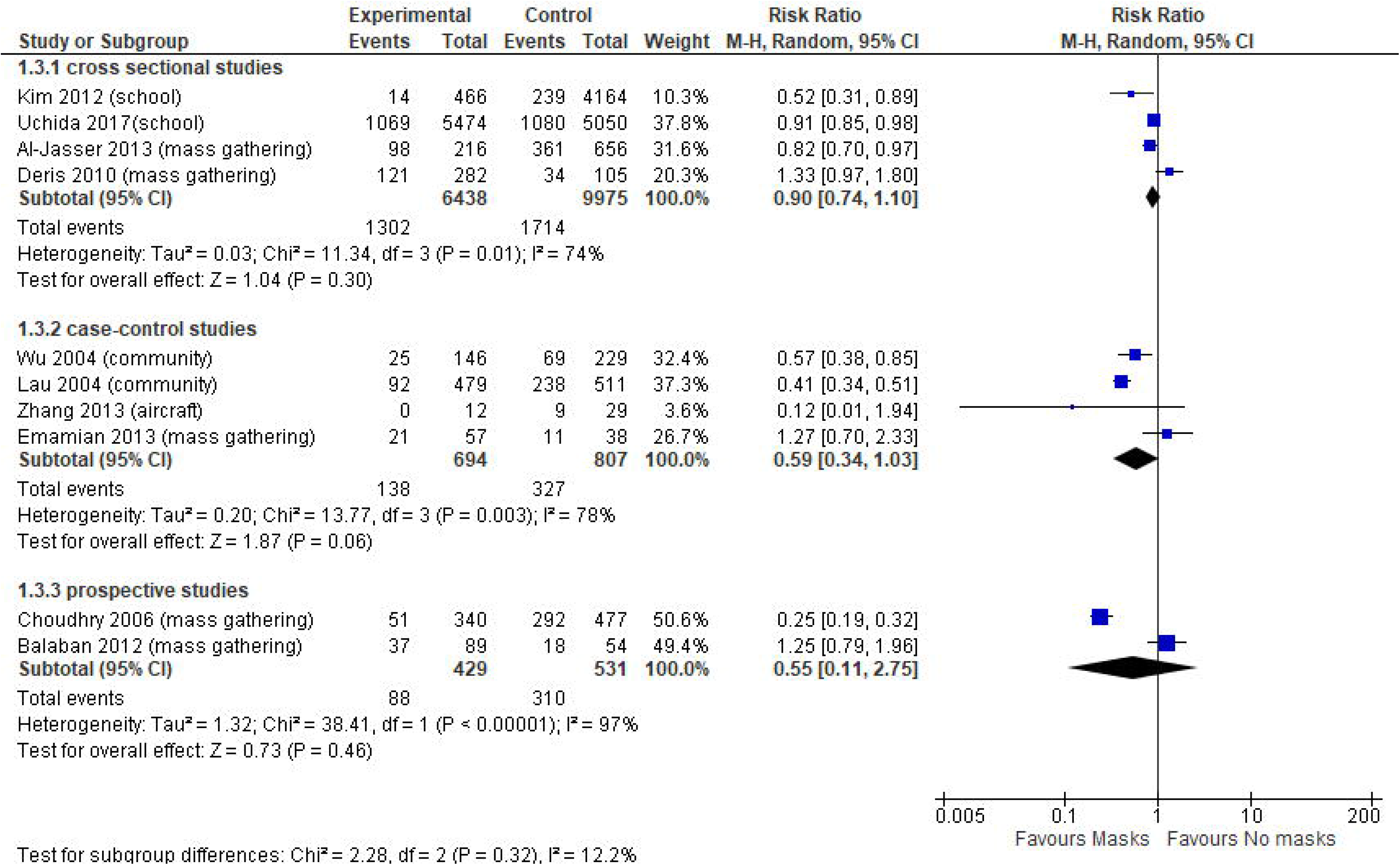
Forest plot of respiratory infection rate in observational studies

Since we found high heterogeneity, we performed sensitivity analysis excluding aircraft and mass gathering studies to determine the robustness of our original analyses and determine whether special settings might have influenced the overall pooled effect. Focusing on studies set in schools, universities and in the general community, the evidence from cross-sectional studies was not statistically significant (OR 0.74, 95%CI 0.43-1.26, I^2^= 76%) (**Appendix 4, Figure S2**); whereas case-control studies (OR 0.46, 95%CI 0.34-0.62, I^2^= 47%) (**Appendix 4, Figure S2**) showed a statistically significant effect in favour of wearing face masks versus not wearing masks, with a more precise overall estimate.

#### Deterministic models

Ten out of 13 studies that deployed mathematical models examined respiratory infection rates (13, 43-53). Across them, three studies (44, 47, 48) reported such an outcome as a pure rate; two papers (46, 52) reported results about the respiratory infections rate as the percentage of cumulative cases; one study (51) reported the number of cumulative cases. The remaining studies did not report intelligible results on respiratory infection rate. Across the above-mentioned studies, the ones that reported the use of N95 masks agree that when at least 50% of the population is wearing a mask the respiratory infection rate can be reduced by a percentage ranging from 80% up to 99%.

The use of facial mask results in a reduction of the respiratory infection rate that is at least of 2.0% in the worst case scenario (52) and up to 99% in the best case scenarios (44, 51, 52). No time-horizon is specified, though. **Appendix 5, Table S9**. The summary of Findings is displayed in **Table 3**.

#### Basic reproduction number (*R*_0_) of viral respiratory infections

##### Deterministic models

Seven out of 13 studies that deployed a mathematical model investigated the *R*_0_ of viral respiratory infections (11, 44-46, 48, 50, 52). Across them, four papers reported explicitly the results (36, 45, 46, 50), two papers only reported a graph (11, 44), and the remaining one did not report accessible data (48). Across these studies, the worst-case scenario was reported in the Brienen et al. 2010 (44): with mask efficacy at 30% and a 20% population coverage, the *R*_0_ reduced from the initial value of 2.0 to just 1.9. On the other hand, the best-case scenario is reported in Chen 2008 (45): with mask efficacy at 95%, the *R*_0_can fall to 0.99 from an initial value of 16.90; however, neither population coverage nor time horizon are reported. **Appendix 5, Table S9** show the description of *R*_0_ of viral respiratory infections across models. The summary of Findings is displayed in **Table 3**.

#### Filtering capacity of masks

Among laboratory experimental studies, seven out of nine studies reported the outcome as filtration rate or face mask protection, including goodness-of-fit and filtration efficiency. Outcomes varied according to the materials used.

In adults, generally filtration rate of household materials had high degree of variation, ranging from 49% to 86% for 0.02 μm exhaled particles (57) and from 3% to 60% for particles in the relevant size range (55).

High degree of variation were also present in surgical masks. One study (55) reported a filtration rate of surgical masks comparable to that of masks made of household materials. Other studies reported the best peformance for surgical masks, filtering from 89% (57) to 95.5%-97% (60) of small particles. Under a pseudo-steady concentration environment, face mask protection on average was found to be 45%, while under expiratory emissions, protection varied from 33% to 100% for fully sealed face mask (58).

Particularly, in paediatrics, penetration of neutralized polydispersed sodium chloride aerosols varied significantly between brands at the highest flow rates, from 15% to 50% (59).

All types of surgical masks provided a relatively stable reduction of aerosol exposure over time, unaffected by duration of wear or type of activity, but with a high degree of individual variation with reductions ranging from 1.1- to 55-fold (average 6-fold), depending on the design of the mask (61). One study compared all types of masks (N95 personal respirators, surgical and home-made masks): surgical masks provided about twice as much protection as home-made masks, with the difference being slightly more marked among adults. N95 personal respirators provided adults with about 50 times as much protection as home-made masks, and 25 times as much protection as surgical masks (62). The summary of Findings is displayed in **Table 3**.

#### Viral load reduction

Three experimental laboratory studies were included (56, 57, 63), of which one study having three arms investigating surgical masks, home-made masks (i.e, cotton mask) or no mask; 2 studies focussed on the comparisons between surgical masks versus home-made masks or no mask (**Appendix 5, Table S10**). According to our PICO, for the SOF GRADE assessment only the comparison between surgical mask versus no mask reporting outcome data was considered (56). This suggested a viral load reduction of 0.25 (0.09 to 0.67) in favour of face mask use (risk difference: 324 fewer x 1000) (**Table 3)**.

## DISCUSSION

This is the first systematic review and meta-analysis comprising evidence based on different research methods and study designs, to address the existing uncertainty about the efficacy and effectiveness of wearing a mask in the community for limiting the spread of COVID-19.

We found very low-certainty evidence that wearing a face mask is associated with a reduced risk of primary infection in RCTs as well as in observational studies. However, the wide confidence intervals affected the statistical significance of the overall estimate. It was not possible to establish the certainty of evidence about mortality, filtering capacity and *R*_0_ whereas viral load was judged to be of very low quality. Our findings indicate (i) a general consensus toward a reduction of deaths when the population mask coverage is near-universal, regardless of mask efficacy; (ii) filtration efficiency depends on the face mask materials, with studies showing high variability. It seems that all types of masks reduce the viral exposure, even though the levels of protection, in terms of reduction of susceptibility to infection in the wearer, are probably lower for some materials (i.e., cloth masks), to the extent that they do not effectively protect against infectious aerosols. Specifically, personal respirators were more efficient than surgical masks, which were more efficient than home-made masks; (iii) in the worst-case scenario with a mask efficacy at 30% and a population coverage at 20%, the *R*_0_ reduced from the initial value of 2.0 to just 1.9; whereas in the best-case scenario, when the mask efficacy is 95%, the *R*_0_ can fall to 0.99 from an initial value of 16.90, even though no population coverage nor time horizon is reported; (iv) wearing versus not wearing a mask is associated with a reduction of viral load of RR 0.25 (95%CI 0.09 to 0.67, based on one experimental laboratory study).

Overall, our findings support the recommendation on using face masks in community settings in a pandemic era: home-made masks, such as those made of teacloths, may confer a significant degree of protection, albeit less strong than surgical masks or N95 personal respirators. Mask efficacy at 95% (N95 personal respirators) seems to be the best scenario, but it is difficult to realize in terms of adherence and costs from a public health perspective. A balanced compromise in the community could be reached with high population coverage using surgical masks (whose mask efficacy is less than 95%), which is easier to implement. Comparing surgical masks to no mask has shown a viral load reduction of a quarter (risk difference: 324 fewer x 1000). Surgical masks were more effective than homemade masks in reducing the number of microorganisms expelled. However, high levels of filtration efficiency have been found among surgical and non-surgical masks, with evidence from all experimental laboratory studies emphasizing the importance of high filtration capacity irrespective of the materials used.

Our findings are in line with results from previous systematic reviews, which however had different aims, population and outcomes. For example, examining the infection rate in pandemic influenza transmission has shown that wearing masks significantly decreased the spread of SARS (OR = 0.32; 95% CI 0.25 to 0.40) (64). Similar finding were found in studies on respiratory virus infections including SARS, H1N1, and COVID-19 in all subgroups, including non-health care worker or non-household contacts (65). One review, investigating the optimum use of different personal protective equipment (face masks, respirators, and eye protection) in community and health-care settings, reported a large reduction in the risk of infection in favour of face mask use (OR 0.15; 95% CI 0.07 to 0.34, RD –14.3%; –15.9 to –10.7; low certainty), with stronger associations with N95 or similar respirators, compared with disposable surgical masks or similar masks (66).

A pragmatic ecologic study involving 49 countries from the European Centre for Disease Prevention and Control (ECDC) investigated the association between face mask use in the community and cumulative number of cases of COVID-19 infection per million inhabitants, discovering that face mask use was negatively associated with number of COVID-19 cases (coef. 326; 95% CI -601 to -51, P=0.021) (67).

The results of this ecological study and of the individual-level studies included in the review are in line with our findings, supporting the use of face masks for reducing the transmission and acquisition of respiratory viral infections in the community.

### Strength and limitations

Our review included experimental laboratory research and mathematical modelling studies to complement observations studies and trials, for obtaining a more complete picture on mortality and viral load reduction, filtering capacity and population coverage which are important factors influencing the *R*_0_. We adopted full methodological rigor within a much shorter time-frame compared to traditional reviews, using enhanced processes. We also critically assessed the risk of bias of included studies (randomized controlled studies and observational studies) and infeasibility of mathematical modelling studies.

Our systematic review has some limitations. We did not investigate the balance of pros and cons of wearing a mask. On one hand, the use of face masks may provide a false sense of security leading to suboptimal physical distancing, poor respiratory etiquette and hand hygiene – and possibly not staying at home when ill. There is a risk that improper removal of the face mask, handling of a contaminated face mask or an increased tendency to touch the face while wearing a mask by healthy persons might actually increase the risk of transmission (10). On the other hand, the fears related to the paradoxical increase of the infectious risk for their improper use are entirely theoretical, based on preconception without real foundation. Education campaigns should be encouraged for assuring proper use (10).

We reported adjusted estimates from two out of three cluster RCTs, because one RCT (32) might have unreliable results due to low usage of face masks in participants: indeed, a low usage of masks was reported in the face mask group, with adherence of only 25% among participants. In contrast, a moderate proportion of participants in the control group (49%) used face masks daily and intermittently. This undermines the reliability of results.

We did not appraise the quality of laboratory experimental studies since we did not find appropriate tools for measuring it. Similarly, for mathematical models we used the unfeasibility appraisal, a proxy of quality assessment, which is more appropriate given the nature of the studies. As for the quantitative-deterministic studies, we acknowledge that such models, especially when SIR-based, do not provide estimates of events, but rather describe what could happen in the future with respect to a predefined set of initial conditions. Namely, they help stakeholders in understanding how the situation could evolve in the future if different actions are adopted today. It follows that pitfalls of such models can be due to mis-specified initial conditions.

With new publications on COVID-19 related prediction models rapidly entering the medical literature, this systematic review cannot be viewed as an up to date list of all currently available prediction models. Furthermore, there were some studies among the ones we have reviewed that were available only as preprints; such studies might actually bring new insights after the peer-reviewing process.

### Challenges and opportunities for public health

The speed of the worldwide spread of the SAR-COV-2 virus, leading to a severe pandemic for which there is no effective treatment or vaccine and limited knowledge on disease behaviour, and the uncertainty regarding the role of asymptomatic individuals in the transmission of the virus, call for Public Health infection prevention and control measures, even in the absence of evidence or in the presence of low quality scientific evidence. A recent systematic review found that, in this pandemic, the proportion of asymptomatic cases ranged from 4% to 41% (68). In this light, universal masking in the community may mitigate the extent of transmission of COVID-19 and may be a necessary adjunctive public health measure (69).

The evidence-based medicine should be used with acumen. The evidence-based GRADE approach suggest that whenever the evidence in favour to the intervention is low but the risks related to the averted implementation could be high, drastic measures can be adopted even in the absence of solid evidence, if the conditions are met (70).

The SARS-COV-2 pandemic is a life-threatening condition to such an extent as to indicate the need of accepting a minimal risk, assuming there is such a risk (i.e., mask costs), of the community intervention (i.e., face mask use), considering the notable benefit of its implementation, even if the evidence-base is of low quality. Deferring these measures, on the other hand, can have a negative effect on health policy decisions. This is called “precautionary principle”: “when human activities may lead to morally unacceptable harm that is scientifically plausible but uncertain, actions shall be taken to avoid or diminish that harm” (71, 72). The evidence, albeit imperfect, in support of the use of masks in this context are justifiable and sufficient in light of this principle. Although evidence-based medicine rightly looks suspiciously at tests of low methodological quality, at the same time it does not completely dismiss them, when circumstances are appropriate, as in this case (8).

And, when it comes to parachuting from a plane that is crashing, you wear it even if no trial has ever shown its effectiveness compared to a control group that launched without (73).

It should be emphasised that the use of face masks in the community should be considered only as a complementary measure and not as a replacement for the core preventive measures that are recommended to reduce community transmission including physical distancing, staying home when ill, teleworking/home working if possible, respiratory etiquette, meticulous hand hygiene and avoiding touching the face, nose, eyes and mouth (10). In conclusion, the use of face masks as single intervention is not sufficient to stop the spread of COVID19 and a full package of the above mentioned interventions is the safest and the most recommended approach.

## Data Availability

Data are available in a public, open access repository. All data generated or analysed during this review are available at https://osf.io/uvjgq/.

## Authors’ contributions

DC, AN carried out the literature search, conducted screenings, extracted data, completed the risk of bias assessment. AF, LI, DD, CR provided a critical revision of the manuscript. SG and GC conceived and drafted the manuscript and performed the statistical analyses. PI conceived and drafted the manuscript, interpreted the data and wrote the discussion. PI is guarantor of the data. CMM and GLT interpreted the data and revised the manuscript for important intellectual content.

## Data Availability Statement

**List of Abbreviations**
aOR: Adjusted Odds Ratio
CI: Confidence Interval
COVID-19: Coronavirus disease 2019
CDC: Centers for Disease Control and Prevention
CHARMS: Checklist for critical Appraisal and data extraction for systematic Reviews of prediction Modelling Studies
ECDC: European Center for Disease Prevention and Control
GRADE: Grades of Recommendation, Assessment, Development and Evaluation
MERS: Middle East respiratory syndrome
MOOSE: Guidelines for Meta-analysis and Systematic reviews of observational studies,
NOS: Newcastle Ottawa scale for non-randomized studies
PRISMA: Preferred reporting items for systematic reviews and meta-analyses
PROBAST: Prediction model Risk Of Bias Assessment Tool
RCT: Randomized Controleld Trials
RR: Risk Ratio
QUADRIAC: QUAntitative-Deterministic models Risk of Infeasibility Assessment Checklist
SARS: severe acute respiratory syndrome
SARS-CoV-2: severe acute respiratory syndrome coronavirus-2
SR: Systematic Reviews
SOF: Summary of Findings
WHO: World Health Organization

